# Applications of artificial intelligence in predicting dengue outbreaks in the face of climate change: a case study along coastal India

**DOI:** 10.1101/2023.01.18.23284134

**Authors:** Krti Tallam, Minh Pham Quang

## Abstract

The climate crisis will have an increasingly profound effect on the global distribution and burden of infectious diseases. Climate-sensitive diseases can serve as critical case studies for assessing public health priorities in the face of epidemics. Preliminary results denote that machine learning-based predictive modeling measures can be successfully applied to understanding environmental disease transmission dynamics. Ultimately, machine learning models can be trained to detect climate-sensitive diseases early, diseases which might represent serious threats to human health, food safety, and economies. We explore how machine learning can serve as a tool for better understanding climate-sensitive diseases, taking dengue dynamics along the Godavari River of coastal India as our case study. We hypothesize that a climate-driven predictive model with controlled calibration can help us understand several of the most critical relationships and climate characteristics of climate-sensitive disease dynamics.

## 1. Introduction

### 1.1 The role of climate in ecosystem dynamics

Climate has always played a critical role in ecosystem dynamics, including in population dynamics and species interactions (1). However, the mechanisms underlying climate variability and its consequences remain poorly understood and infrequently field-tested (2). This is particularly true because climate effects often appear context specific (3). Multiple climate variables can act synergistically, individual climate variables can affect multiple aspects of ecosystem dynamics, and impacts can often be nonlinear, challenging to predict in the face of the climate crisis, and presented across a gradient of important conditions (4). Therefore, surprisingly, while climate is known to be one of the most prominent influencers of ecological processes, its dynamical and directional effects on ecosystems are often poorly understood and challenging to predict. This has become increasingly true against the backdrop of the climate crisis and increasing anthropogenic stressors and especially so for coastal communities (5).

### 1.2 Climate-sensitive diseases

Climate-sensitive diseases provide a critical case study to model and measure whether climate-sensitive traits simulated in models can help us more accurately assess the multitude of dynamics observed in the field. For mosquito-borne diseases in particular, rising average temperatures have often led to geographic range expansions of disease vectors, decreased incubation periods of some pathogens, and increased rates of contact of some mosquito species that prey on humans (6). Recent knowledge provides evidence that many ranges of mosquito-borne diseases will expand dramatically in response to the climate crisis (7). Models have also helped scientists better understand how average sea levels can also influence the density of salinity-tolerant climate-sensitive pathogens, namely, mosquitos, along coastlines (8). Statistical simulations have helped point science to better quantify the influence of increasingly warmer climates on the hydrological cycle, a phenomenon which has been in part discussed in the context of the extreme weather events observed increasingly frequently over the last few decades (9).

### 1.3 Mosquito-borne diseases

The impacts of climate change on mosquito-borne diseases are intensified by shifting economic, demographic, and social factors making it even more necessary to better model how environmental patterns may be shifting in areas that are increasingly prone to climate-sensitive infectious diseases (10). Even going a few decades back, Johansson et al. and Moore et al. discuss evidence of unpredictable temperature and precipitation influencing the transmission and abundance potential of *Aedes aegypti* in Puerto Rico (11,6). Suggestive relationships have also been discovered between coastal dengue outbreaks and sea surface temperatures in places such as New Caledonia and Mexico (12–14). The transmission of mosquito-borne, viral, climate-sensitive diseases such as dengue occur along a spectrum, from low levels of year-round endemic transmission (15) to larger-scale period or interannual outbreaks (16). We hypothesize that the range of dynamics of mosquito-borne disease dynamics are a result of regional or seasonal differences in climate, where the magnitude or direction of climate effects on mosquito-borne vectors differ (17–19).

### 1.4 Prediction and intervention of disease dynamics along coastlines

Systematically understanding the drivers of climate-sensitive mosquito-borne disease dynamics will significantly support two core scientific outcomes: the prediction of the climate crisis’ effect on disease dynamics, especially along coastlines, and the intervention and management strategies required to prevent detrimental outbreaks (20). For instance, predictive models of dengue dynamics that can capture small-scale mosquito population dynamics along coastal India can be applied to more realistic projections for outbreaks dynamics across other tropical coastal regions (21,22).

Despite the vitality of predictive approaches, validation with vector and disease data remain limited given the challenges of obtaining and processing such data. Thus, it raises the question of necessity in that scientists must assess which disease variables a model should capture based on model parameterization. This must be either scalable or independent from the transmission system researchers aim to predict. Since disease dynamics cannot be studied in the context of every possible permutation of climate parameters, transmission setting, or scale, understanding the extent to which models derived from fundamental science or modeling-based traits will be critical for translational and scalable modeling of future climate-sensitive mosquito-borne disease dynamics (23,24). We hypothesize that a climate-driven predictive model with controlled calibration can help us understand several of the most critical relationships and climate characteristics of climate-sensitive disease dynamics.

### 1.5 Dengue outbreaks along coastlines

In this work, we test the extent to which climate-driven mosquito traits drive disease dynamics across a coastal region of eastern India, given the climate dynamics of the Bay of Bengal and the opportunity to characterize additional climatological and ecological factors that may mediate the effects of climate on dengue disease dynamics. Given the ecology of the primary disease vector of dengue, *Aedes aegypti*, and the fact that they are anthropophilic, their traits are important for the transmission of dengue. These traits include their reproduction, survival, development, biting date, and extrinsic incubation periods. Studies have demonstrated that these traits are heavily temperature-dependent with an intermediate thermal optimum (25,26). Humidity has been positively associated with mosquito survival because of the high surface area to volume ratio of mosquitos exposed to desiccation (27). Standing or stagnating water resulting from rainfall also provides ideal pupal and larval habitats for mosquitos (28). The relationship is also challenging because heavy rainfall or extreme weather events off the Bay of Bengal (i.e., cyclones) can also flush away *Ae. aegypti* breeding habitats. Recent studies have demonstrated that models incorporating temperature-based climate traits can provide insights on some critical features of dengue disease dynamics (29).

### 1.6 Historical understanding of dengue and climate in India

Previous studies have attempted to understand the relationship between climate and the presence of dengue in India by means of statistical and mathematical models. These techniques have included Poisson regression (30), Naïve Bayes and multivariable regression (31), Bayesian models such as spatial autoregressive (SAR) models or conditional autoregressive (CAR) models (Mudele et al., 2021), autoregressive integrated moving average (ARIMA) models (32), generalized linear models (33), Extreme Gradient Boosting and rule-based classification (34), and other types of mathematical modeling (35). The above methods have focused on multiple parameters influencing dengue and on the discovery of the climate variables that most influence dengue disease dynamics (36).

From our literature review, we observed the establishment of many statistical and mathematical models, which address stochastic, mechanistic, and deterministic dengue disease models, among others. However, there is still limited research probing the predictive modeling of the complexities of dengue disease models with the backdrop of climatically critical variables. One tool that has recently surfaced as an emerging modeling strategy is machine learning modeling for predictive dengue disease dynamics (37). While there exists research into the effects of dengue dynamics in relation to climate, these are often backed by traditional statistical models. Machine learning tools offer a critical opportunity on capitalize on greater predictive ability. This can expand the literature to include state-of-the-art models for assessing dengue disease dynamics. In this study, we used advanced machine learning techniques to assess the impact of hydroclimatic and extreme weather climate parameters on dengue disease dynamics and outbreak predictions.

## 2. Materials and Methods

### 2.1. Data development and visualization

#### 2.1.1 Data cleaning and augmentation

Our data were hand-collected on the ground between 2019-2020, in Andhra Pradesh, India. Along with historic data collected at those points in time as well, the dataset spans 2016-2020 with weekly frequency. The months of January to March were not examined in this study due to data quality and collection issues. The following climatic parameters were used in our model: windspeed, pH level, dissolved oxygen, temperature, total number of extreme weather (measured via cyclonic) events that took place, rainfall, precipitation. They were chosen based on previous literature and on hypothesized relationships to dengue fever. The climatic data were collected from the surrounding area of the Godavari River with primary focus in subsections of the rivers in Andhra Pradesh, India. To evaluate the accuracy of models in determining the relationship between these variables and dengue disease dynamics, we used dengue case counts as our predictor benchmarks for both regression and classification tasks (explained further in Section 2.2).

#### 2.1.2 Model parameters

##### Wind speed

The model parameter “wind speed” was added based on the provided wind speed data. In the original dataset, wind speed was represented by weekly occurrence and measurements. After several model iterations, it became apparent that those weekly changes were unsuitable for the nature of this long-term dataset. Therefore, when a cyclone was documented, the full month of its occurrence was labeled with the respective wind speed. For months during the dataset that had multiple cyclones occur, 2-week or 3-week intervals were labeled with the wind speed of the cyclone. For months where no cyclones occurred, the default value was set to be a normal wind speed of 20 kilometers / hour. The methodology has potential for further fine-tuning during future model iterations.

##### pH and dissolved oxygen levels

pH and dissolved oxygen levels were obtained using data from a subsection of River Godavari at Rajahmundry (16.714456°, 82.331298°). The measurements were originally obtained in monthly intervals. For the purposes of our model, we had to down-sample the data to weekly intervals. Hence, in the final dataset, the weekly data points are grouped by month. The data was obtained near the outtakes of water treatment stations through the Water Resource Board of Andhra Pradesh.

##### Dengue counts

Dengue fever is a highly seasonal disease and, for our purposes, is spread through the bite of an *Aedes aegypti* (*Ae. aegypti* hereafter) mosquito. Being a mosquito-borne disease, dengue demonstrates high correlation to the mosquito population in the region (38). *Ae. aegypti* populations thrive within conditions created by monsoons (humid, damp, brackish waters). As such, increased dengue cases, and more frequent dengue outbreaks, usually occur during the monsoon season.

##### Additional Model Parameters

For model features not mentioned above, they were collected by the lead author and sourced during the time spent in the region of Andhra Pradesh.

#### 2.1.3 Time series analysis

A high degree of seasonality was exhibited across all features. For all climatic variables, we observed a yearly spike during the months of April through July. The rapid spike in this period for all variables indicates active environmental activity in the form of increased rainfall and large numbers of storms and cyclones. These are indicative of typical tropical climates, especially in the monsoon season, although with climate change have increased severely in the last decade. Moreover, the number of dengue cases also shows a spike in these months, which suggests there is a correlation between extreme weather events and dengue outbreak risk in the region. This can be further interpreted as allowing for an increased exposure to *Ae. aegypti* mosquitoes due to the ideal breeding conditions facilitated by climate conditions (damp, humid, and stagnant water bodies). Our models show that this often leads to an overall increased in mosquito population in the region and increased risk of dengue cases [**Figure 1**].

**Fig 1.**
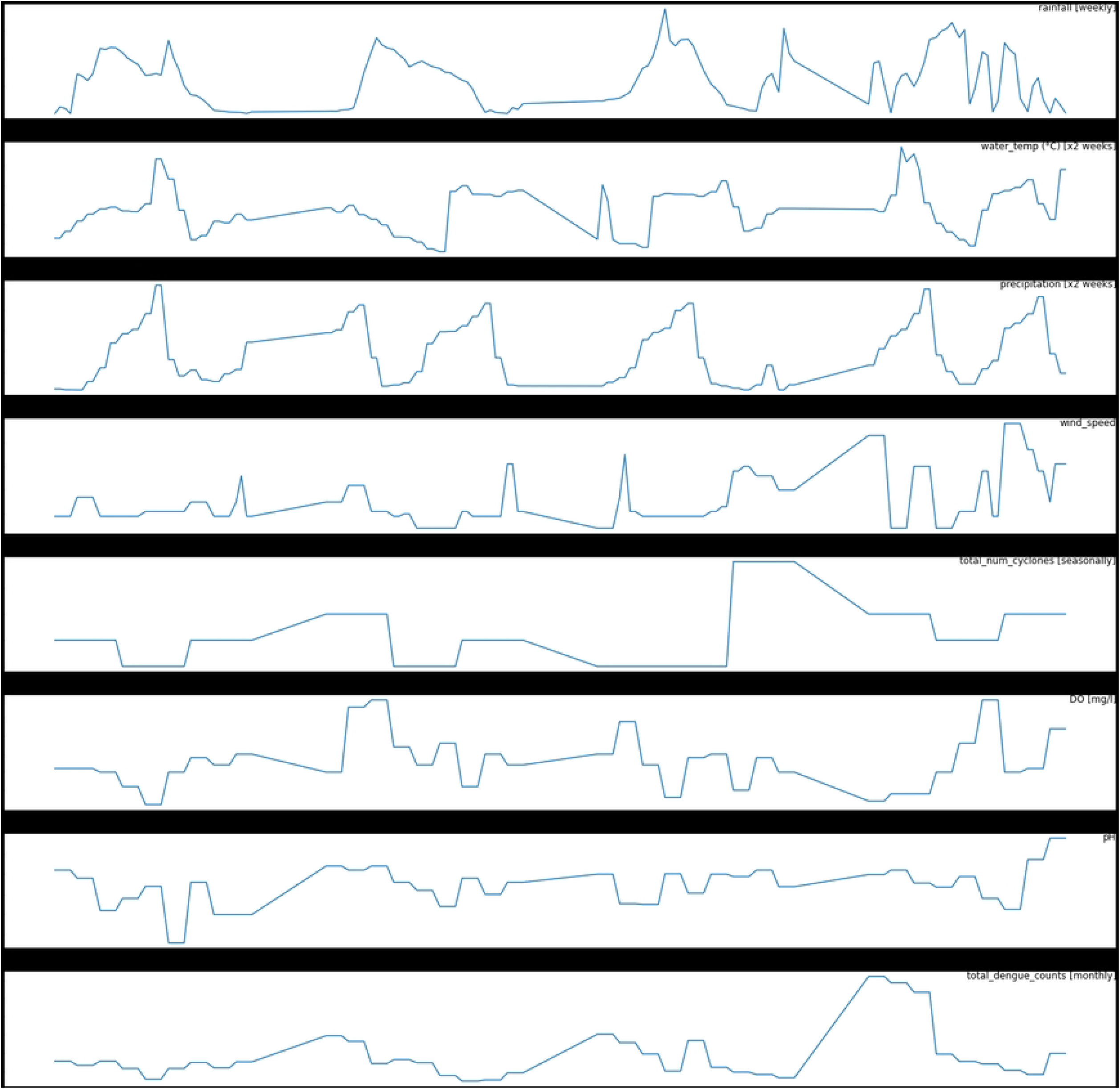
Our time series analysis of the variables used in our study.

By observing the trend lines more closely, there exists a strong correlation between rainfall, precipitation, water temperature and dengue counts. It is worth noting how dengue counts demonstrate a lag of 2-4 weeks after a spike in rainfall and precipitation [**Figure 1**].

In 2019, we note a particularly abnormal increase in the number of dengue cases, with approximately 40 percent more cases than the average number of cases in previous years. The outlier years in dengue cases were also followed by the highest spike in other climatic environmental variables. This could indicate that there was a heavy dengue outbreak in the region that was likely, at least in significant part, influenced by climate. Given this, we broke down a correlation matrix to observe the individual relationships between variables [**Figure 2**].

**Fig 2.**
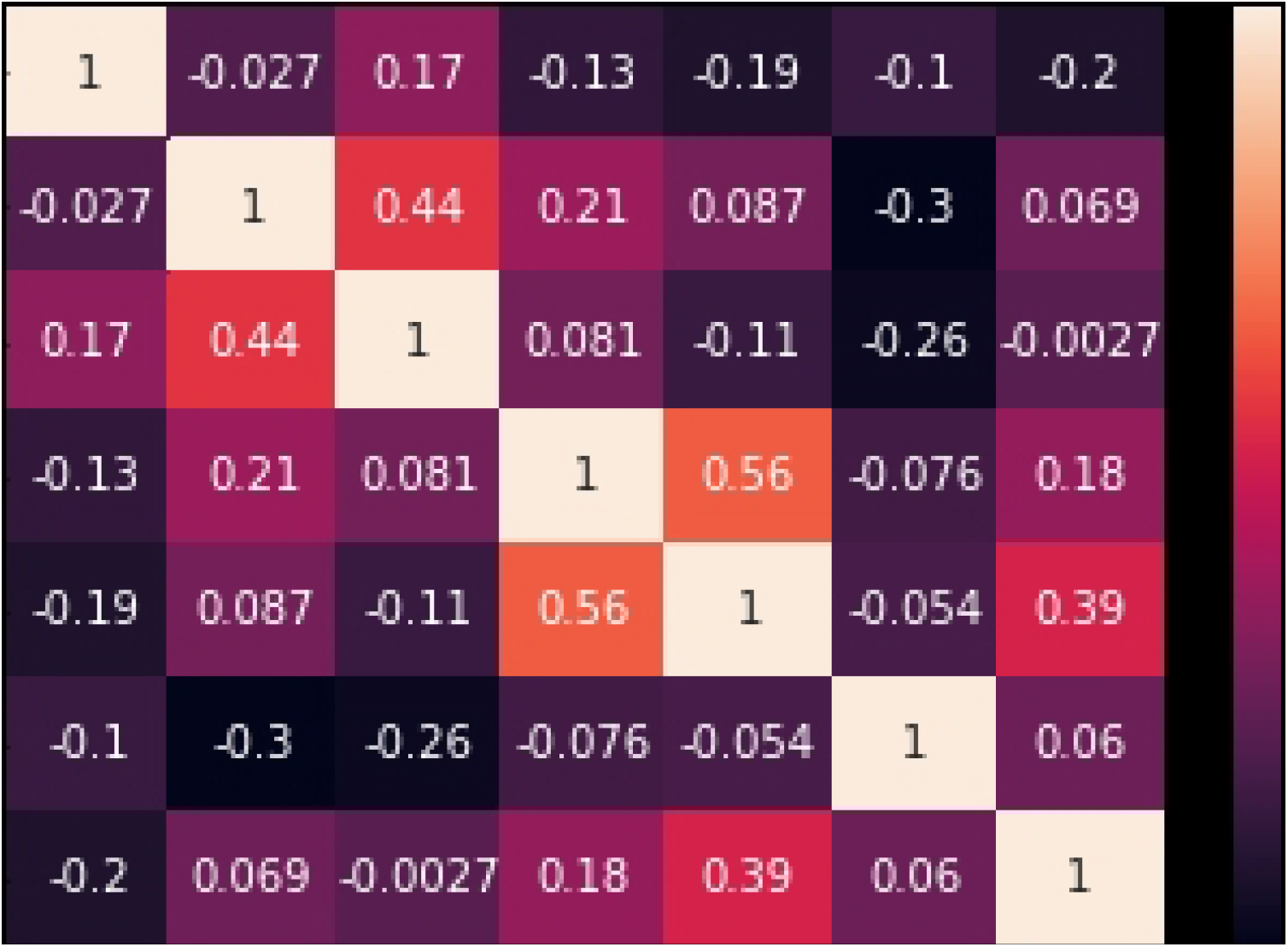
The correlation matrix of all climatic variables in the study.

#### 2.1.4 Correlation analysis

We note the strong positive correlation between number of cyclones and wind speed [**Figure 3**].

**Fig 3.**
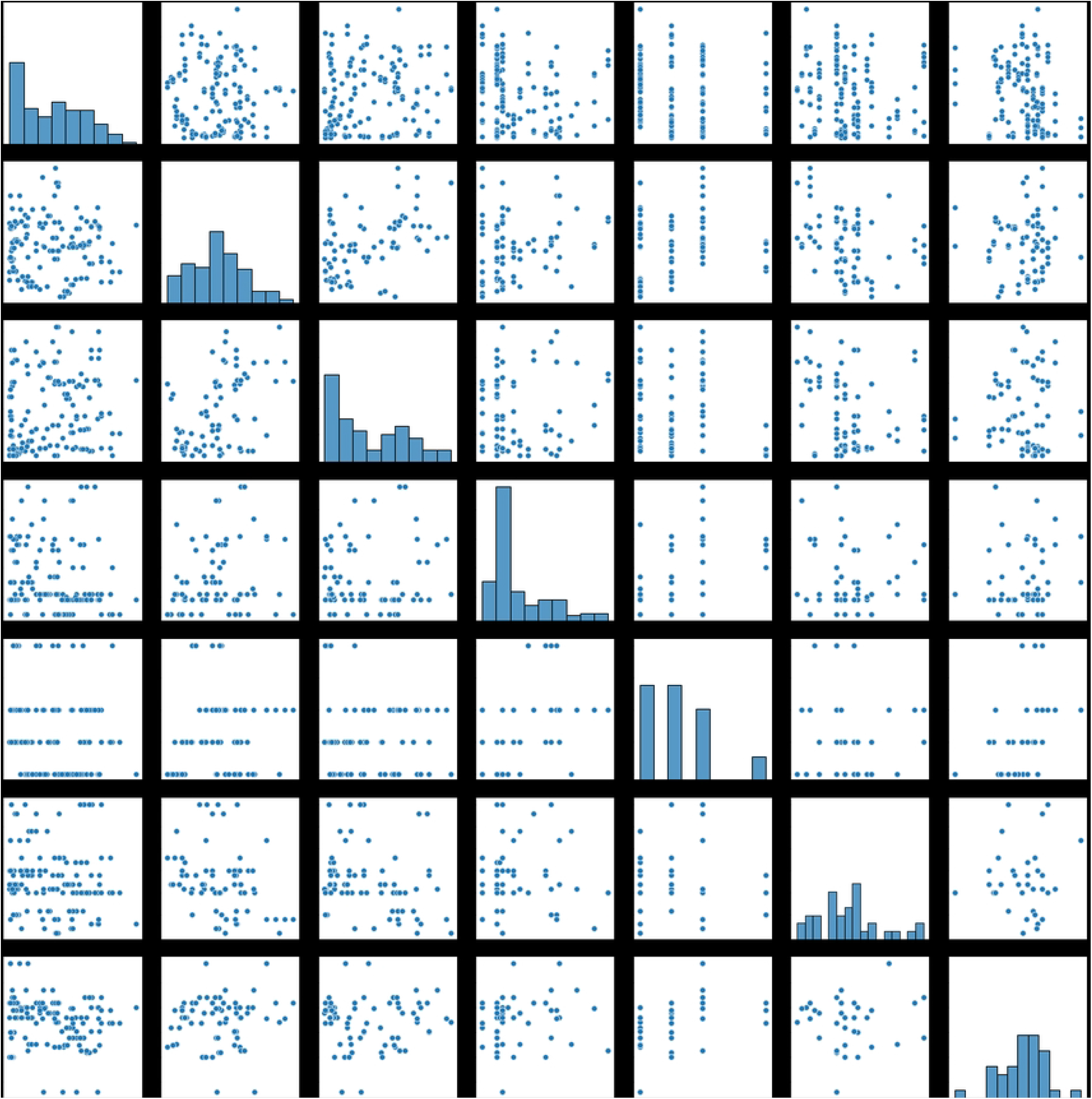
Correlogram of all climatic variables in the study.

This is logical given the that cyclonic activities would cause fluctuation in wind speed data. We also observe the strong positive correlation between precipitation and water temperature, and a strong positive correlation between total number of cyclones and ocean water pH levels. There is a moderate positive correlation between wind speed and water temperature. There exists a strong negative correlation between precipitation and dissolved oxygen levels.

For the other features of the model, there was a relatively weak correlation between each variable, indicating that each of those variables showed greater independence from one another. However, we use this to our advantage as it can provide clearer insight into what influenced dengue counts and resulting outbreaks in the region. This is critical for scalability and transferability of our dengue model because it will produce generalized results that are not biased towards a single climatic feature.

### 2.2 Machine learning models: data augmentation

#### 2.2.1 Experimental Setup and Software

Both classification and regression methods were performed during analysis. In this study, all data processing and analysis was conducted using Python 3.9 on Google Colab. To produce results, NumPy, scikit-learn, Keras libraries were employed in the cleaning and model development process.

#### 2.2.2 Balancing the dataset

We observed a relatively imbalanced dataset. Hence, we resampled our dataset using a Python scikit-learn class known as Synthetic Minority Oversampling Technique (SMOTE), which acts as a data transform object for classification datasets that observe imbalance in their datasets (39). SMOTE duplicates examples in the minority class and new examples can be synthesized from the existing examples. We note the newly generated dataset has a near balanced number of samples, with 288 samples in total used for both the training and testing datasets.

#### 2.2.3 Data Preprocessing

Given the data collection process, normalization of variables is necessary. As different variables were measured on different scales such as mm for rainfall and km/h for cyclone speeds, these differences may influence the weighting of each parameter so that the models are heavily influenced by variables maximum values, creating bias. Moreover, normalization helps models detect hidden pattern in data more easily as they helps detect the minor variations in data such as in pH levels more easily. We normalize all our variables using Min-Max normalization on our dataset to ensure each data point was scaled to the same range from 0 to 1. Min-max normalization is a common way to normalize data, where for every feature, the minimum value of that feature gets transformed to 0, the maximum value gets transformed into a 1, and every other value is transformed into a decimal between the scale of 0 and 1. The Min-Max normalization function is defined as equation:

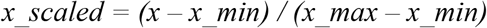

Where x is the variable being scaled, x_max is the maximum value x over all observation of the variable, x_min is the minimum value x over all observation of the variable and x_scaled is our result.

#### 2.2.4 Time-lagged dataset

For both of our classification and regression tasks, a 4-week time-lagged dataset was used. The rationale behind choosing such a timeframe was both from a machine learning and ecology standpoint. We note that after trialing different time spans for the lagged dataset, the 4-week time lagged dataset offered the best performance for the models while it did not cause the models to overfit due to having many parameters the models were inputted. From an ecology standpoint, 4-week time lag would capture the growth cycle of the *Ae. aegypti* mosquito after laying eggs in brackish water. Dengue transmission requires multiple developmental processes to occur in the mosquito and parasite, resulting in time lags that vary with temperature. Time lags also arise between climate and dengue transmission because temperature affects mosquito development rates in maturing stages, as well as during the oviposition cycle and during pathogen extrinsic incubation periods (28).

### 2.3. Machine Learning Models

#### 2.3.1 Regression Methodology

We aimed to produce predictive forecasting curves of dengue counts using long short-term memory networks (LSTM) models. LSTM models are used in deep learning and machine learning as a variety of recurrent neural networks (RNNs) capable of learning long-term dependencies, especially in sequence prediction problems. We used a time-lagged dataset (explained in Section 2.2). The dataset was built using the 4-week lag time frame, as it turned out to be most adequate in representing seasonality of dengue transmission, while not still preventing the model from overfitting, which we discovered from both our own data and other studies (25). We employed the Root Mean Square Error (RMSE) to calculate the prediction errors of the model. We trained the model on 100 epochs.

With initial experimentation, the models indicated a high degree of overfitting. This is seen through validation scores not decreasing in accordance with a decrease in training score. In fact, it would increase with more training epochs. This is common in machine learning tasks due to various reason including noisy data or small datasets.

To amend the situation, we added dropout layers to help decrease overfitting. We also designed multi-layered LSTM model, with 3-layered setup so that each LSTM is independently trained from each other so that it could generalize more and learn better. Parametric Rectified Linear Unit (PRelu) as an activation function is used due to its ability to prevent overfitting and because it is commonly used in similar time series models (40).

#### 2.3.2 Model Metrics

##### RMSE (Root mean squared error)

RMSE is one of the most used measures for evaluating the quality of predictions. RMSE is equal to the square root of the average of squared residuals. The RMSE penalizes heavily on large errors as the residuals are squared before taking the average. In the case of our model, it is particularly useful when large errors are undesirable as it helps model to produce better predictions (41). High reliability is particularly desirable as dengue forecasting is used for policy changes, which requires accurate information to maximize effectiveness.

#### 2.3.3 Models

##### Recurrent Neural Network (RNN) Models

RNN is a type of neural network that handles previous outputs as inputs, factoring in historical information into the model. As a neural network model, they provide researchers the ability the ability to extract relationship between complex climatic, hydrological variables and dengue fever risk. The RNN model allowed for greater predictive ability given the highly seasonal nature of dengue fever, as it is developed to handle timeseries data (Feng et al, 2021). In this study, we opted to only develop an LSTM model, an extension of RNN model, as RNN models face vanishing gradient problems. This influences their ability to work with long-term historical data. That is, the influence of the earlier inputs ‘disappears’ when the hidden layer is overwritten with new inputs, this causes the network to ‘forget’, which hampers the ability of an RNN to handle higher long-term dependencies.

##### Long-Short Term Memory (LSTM) Models

LSTM is a type of RNN developed to overcome the vanishing gradient problem faced in traditional RNN models (Hochreiter & Schmidhuber, 1997). LSTM models learn long-term dependencies and extract and retain information from a much longer timeframe compared to RNN. LSTM models achieve this with the introduction of memory blocks eliminating the vanishing gradient problem (Sundermeyer, 2012). In doing so, LSTM models better captured the relationship between historical trends of dengue cases and the complex climatic conditions for the purposes of our model.

##### LSTM with Attention

LSTM models can also lose important information when passing information across multiple sequence steps, Moreover, LSTM models primarily work with fixed-length time sequences without special consideration of the different time of the year, such as pre-monsoon or monsoon season which influences the importance of different climatic variables. LSTM with Attention is an extension of LSTM models which introduces attention mechanism which is a method to deal to with non-fixed length sequences of time and allows LSTM models to selectively pay attention to the current input. This ultimately enabled a stronger understanding of our timeseries data. In our study, LSTM and LSTM with Attention were also compared to determine the suitability of each model in predicting number of dengue cases (Shook et al, 2021).

#### 2.3.4 Classification Methodology

Instead of using binary classification to identify which weeks are more prone to dengue case increases, we decided to use multiclass labels to better represent dengue risks of a specific week or set of weeks. The categories were: low risk, medium risk, and high risk (with corresponding labels of 0, 1, and 2).

We chose to use dengue prediction counts of specific months to categorize our data. Low risk weeks were weeks with lower dengue counts than the mean dengue counts, medium risk weeks were at most ¼ standard deviation higher than the mean dengue counts, and high-risk weeks were weeks with more than ¼ higher standard deviation higher dengue counts than the mean dengue counts.

The threshold was defined in the following ways based on prior research on the matter (Pham et al, 2022). We also considered the outlier weeks where extreme numbers of dengue cases were presented (i.e., counts that were several times the mean).

#### 2.3.5 Metrics

##### Accuracy

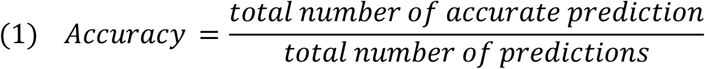

Equation (1) demonstrates how well a classifier model will perform in our multiclass classification problem. Its calculation provides a clear indicator to the performance of a model as it computes the total number of correct predictions divided by the total number of predictions made. Accuracy is the most often used metrics in classification problems.

#### 2.3.6 Models

##### SVM

Support Vector Machine (SVM) models are methods designed for classification and regression problems. The algorithm performs predictions by establishing the best hyperplane or hyperplanes that minimize the loss function over the feature space. An SVM would determine the hyperplane by calculating the maximal margin of the classes which is the maximum distance of the nearest element to the hyperplane for each class, with the nearest element of each class to the hyperplane being called support vector as the affects the orientation of an SVM. However, this can only be effectively done if the relationship between variables for each feature are linear, for non-linear inputs this problem is resolved via a kernel. A kernel is a function which allows the model to project the feature space into higher dimension until each class is linearly separable. The choice for kernel function in this article was radial basis function based on previous related literature.

##### KNN

K-Nearest-Neighbor (KNN) is used for a variety of problems in ML. KNN is a stable deterministic algorithm which produces the same result across trial runs and is computationally efficient, only needing to calculate the relationship of an observation and “k” of its nearest neighbor. K here is defined as the number of neighbors being weighted in calculating the cost of each sample point. Here k = 7 was used to generate the most optimal result (Gauahr et al, 2021). The algorithm works by calculating the distance functions between each of the nearest neighbors and then classifying them based on a majority vote based on distance weights.

##### Random Forest

Random forest models are ensemble models that compute results based on the aggregate of multiple random decision tree classifiers. The decision trees in random forest are generated from a random subset of features, compared to normal decision trees which are generated from the entire dataset. The decision trees in a random forest model are created to have low correlation between one another, which helps the random forest model perform better than decision tree models (42). Random forest models are particularly useful in dealing with predicting dengue fever case variability, as they can deal with numerous independent variables efficiently. Another benefit is that when multiclass classification is employed, random forest models performed well given that they are designed to handle such problems without much extension being required of the base model.

## 3. Results

### 3.1. Results of regression models

From **Table 1**, we can see that the LSTM captures the trend of cases increasing, indicating the high spread of dengue cases during in monsoon season. It was interesting to discover that each model provides similar regression curve with the LSTM with Attention model even providing a worse RMSE score in the validation dataset compared to the standard LSTM model. A potential explanation can be attributed to the fact that the time frame used was limited using only 4 years, as such the added complexity of LSTM with Attention hinders its predictive ability. The dataset is also limited for a machine learning algorithm. It was also found that the upward and downward trends simulated and forecasted by the LSTM model provides a reasonable approximation to the reported points, especially for the identification of high incidence peaks, which supports the results where LSTM can better capture the trends in dengue cases. Another noteworthy feature is that models can accurately capture the high jump in dengue cases at 120 weeks after the start of the dataset, the outlier high of dengue cases we see here can be attributed as a major dengue fever outbreak in the region and the regression results indicate the models can follow such trend accurately, proving their usage in outbreak detection. This study serves as support that deep learning models such as LSTM can be beneficial in forecasting dengue outbreak, allowing for authorities to better prevent dengue cases.

**Table 1.** RMSE scores of all regression models.

| Model | Train Score | Test Score |
| --- | --- | --- |
| LSTM | 0.04 | 0.12 |
| LSTM With<br>Attention | 0.04 | 0.17 |

### 3.2. Result of the classification models

Overall, all models show promising classification results [**Figure 4**]. The accuracy of all models indicated were above 90 percent, with most misclassifications being in low risk and medium risk weeks with high reliability. It means that the parameters selected on the time-lagged dataset have contributed information for the models to analyze and improve from. In the SVM models, there were the most totaling 3 misclassifications on low dengue risk week prediction, indicating that the model would have some difficulty differentiating between low dengue risk week and medium risk. An explanation for this would be due the thresholding value as it medium risk prediction has a smaller range compared to high risk and low risk value causing models to make wrong predictions more frequently, which indicates that current models still lack the ability to discern smaller variation in changes between climatic variables that causes changes between each type of dengue risk classification. Moreover, SVM models do not support multiclass classification natively which could mean that the extension to handle multiclass labels which hamper to performance of SVM models.

**Fig 4.**
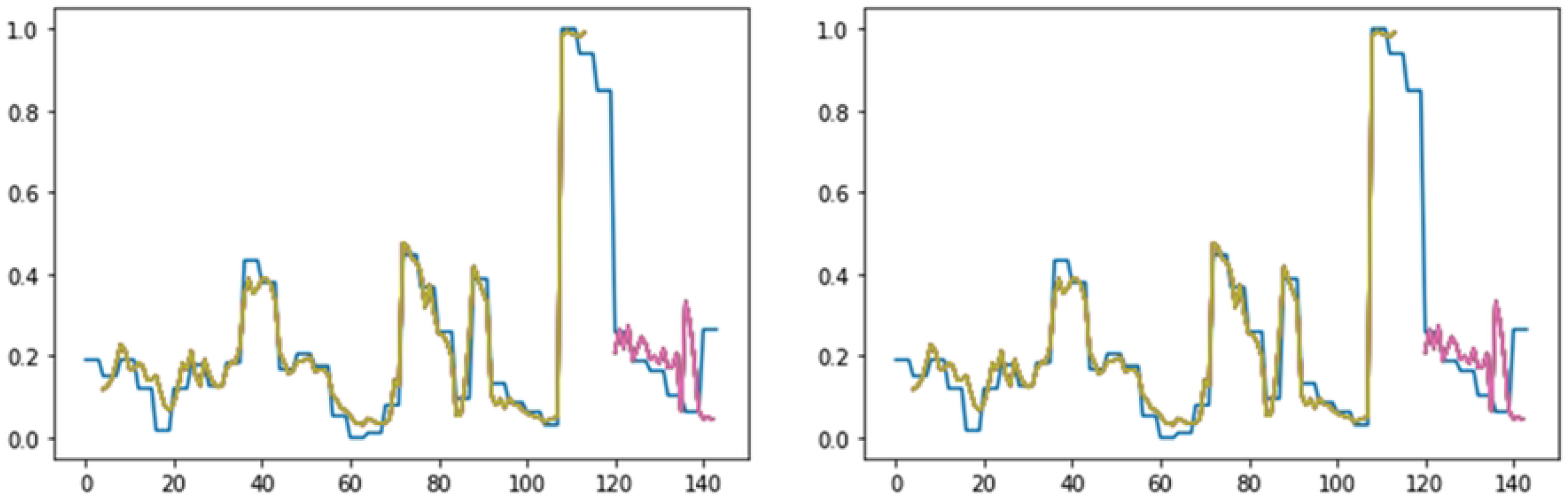
The regression curve of the LSTM and LSTM with Attention.

Comparatively, both KNN and Random Forest models performed much better in classifying both low risk and medium risk weeks [**Figure 5**]. All models show capacity to predict high risk dengue week. This can be attributed to extreme weather events influencing climatic hyperparameters, allowing for models to better detect high risk weeks. From the current results, it can determine that the models employed can generate accurate predictions with some models performing at 97 percent prediction on a time-lagged climatic dataset.

**Fig 5.**
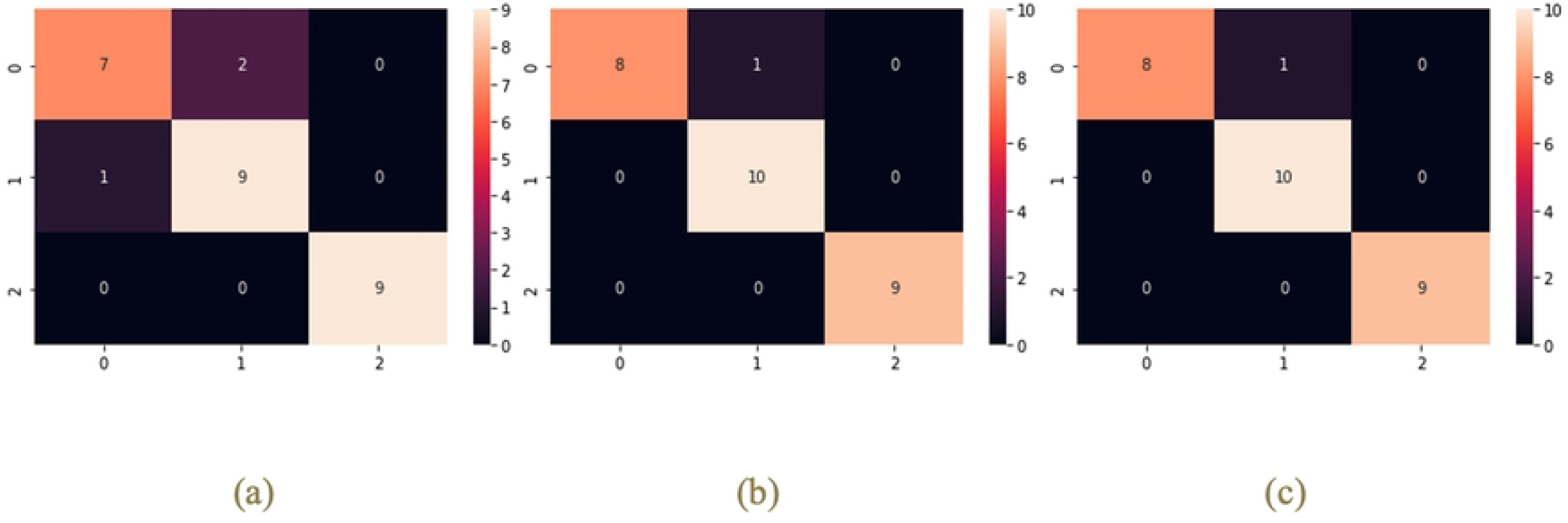
The confusion matrices detailing the prediction result of each model: (a) Confusion Matrix of SVM. (b) Confusion Matrix of KNN. (c) Confusion Matrix of Random Forest.

## 4. Discussion

### 4.1 Summary of our results

The purpose of this study was to dynamically model dengue transmission dynamics in tropical India by the contributing temperature-dependent entomological parameters of *Aedes aegypti*. Our study provides a dynamic model of dengue transmission along the Godavari River district of eastern coastal India. We incorporate seven climate parameters into a predictive transmission model and consider the sensitivity analysis of dengue cases. The benefits of this model are that they can present how dengue transmission dynamics are influenced by factors such as extreme weather events (i.e., cyclones). This model only considered limited climate and dengue parameters to provide a proof-of-concept for more complex models in the future. Moreover, our models operate without examining less accessible satellite imagery and GIS data. As these models are operate with only numerical 1-d data points, it provides a low barriers of entry methodology for integrating ML models and further research in dengue forecasting.

To demonstrate the feasibility of the models, we explored dengue modeling through both regression and classification lenses, to showcase the many potential development further research can dive deeper in the field. In regression, both LSTM and LSTM with Attention models were explored. These models produced extremely desirable results in determining the dengue cases in the regions with RMSE below 0.2. These scores are comparable to similar models explored in different regions (Pham et al., 2022). In classification, it is determined that SVM and Random Forest models provide the most accurate result and most suited to identifying and differentiating between low risk and high-risk months and dealing with multiclass labels. This is due to the nature of these algorithms as they are developed to natively support the classifying multiclass labels.

The results of the study provide interesting insights into dengue fever forecast. Through feature ranking, it was determined that temperature, windspeed and rainfall are variables of the most important overall to dengue prediction models. Their value to models can be easily explained through observing the extreme weather events in India and their effects on *Aedes aegypti* population in the region. Cyclones often introduces environments where mosquitos’ population thrives, therefore increasing the spread of mosquito-borne diseases. Moreover, these models work well with classifying outliers’ dengue cases week. This means that ML models can handle the task of determining outbreak time periods as they have shown the ability to both capture and predict radical changes in dengue fever case counts and accurately classifying high dengue risk week.

### 4.2 Applications of Machine Learning Models in dengue outbreak forecasting

Climatic variables provide suitable indicators for machine learning models to take advantage of its predictive ability (43). Given the increasingly unpredictable nature of climate change, traditional statistical models and master systems that rely heavily upon human calibration and expert knowledge are increasingly unsuitable and inflexible to changes in environmental systems (44). ML models provide a solution in the face of a rapidly changing environment. As such, these models seek to determine the indirect relationships between climatic variables and climate-sensitive mosquito-borne disease dynamics without the need for heavy human monitoring. This minimizes the biases influencing predictive models, while enabling such models to rapidly adapt to changing ecological systems through continuous updated data and replenished training. Moreover, ML models can extend well to different conditioning factors without the need to redesign model architecture (45).

ML models can extract information on much more extensive set of inputs as compared to mathematical models and statistical models. A diverse set of inputs, including hydrological and extreme weather data, helps models generalize better forecast dengue outbreaks. This is because they can better simulate ecological changes that influence *Ae. aegypti* populations (25). While more traditional models operate on more fixed inputs and required extensive efforts to extend their feature space, ML models can easily work with multiple conditioning factors without much additional extension to the base model, enabling for better development time and adaptability.

ML can provide prediction results on imperfect or missing information, a necessity given that environmental data is often not extensively available, even more so in rural or under-resourced regions. This enables better reach and understanding of epidemic diseases as it is often in these regions that these outbreaks are most widespread and most destructive, given a lack of medical resources and disease monitoring capabilities. Delayed dengue case reporting or incomplete meteorological data can influence the effectiveness of ML models; however, such a tradeoff is acceptable in real-world implementation of the ML models presented in this study, as more traditional models often cannot forecast on missing data.

### 4.3 Dengue and climate change

It is established that we are in a increasing climate crisis and that will influence climate-sensitive mosquito-borne disease dynamics in different capacities across the world (8). There are many mosquito vectors that transmit important human parasitic and arboviral diseases, but dengue remains the most common human arboviral disease. It is reported to have a prevalence of 50 million cases in more than 100 countries, with about 500,000 persons requiring hospitalization each year for dengue hemorrhagic fever/dengue shock syndrome that has overall case fatality rate of 2.5 percent (46). The mounting evidence around climate-disease relationships raises many important issues about the potential effects of global climate changes on the transmission of dengue.

Although there is growing statistical evidence indicating that dengue outbreaks are associated with temperature, rainfall, and humidity, few studies have examined this relationship along coastlines and with the inclusion of extreme weather events, including both a critical spatial component (47) and a critical climate event worth examining (37). The authors have identified that other variables such as river levels are also included along with climate variables (48), however, these studies are out of the study scope. From our study, it is critical to include a variety of geographic topographies and regions in dengue transmission dynamics models moving forward, but it is also critical to model which climate variables are the best predictors of dengue transmission. The research in this study indicates that cyclones are significant contributors to dengue prediction models and must be included going forward.

### 4.4 Dengue and Coastlines

Dengue demonstrated an uneven distribution along the Godavari River of Andhra Pradesh and expressed differently on the spatial gradient from coast to inland. The temporal trends for coastal dengue showed cyclical trends with 3 to 5-week lags between cyclonic events and a peak in dengue counts. This is interesting because inland, the inland lag time has been estimated to be an average of up to two months (25) but coastal lags appear to be much smaller time frames in this case. Our findings demonstrate that coastal dengue transmission dynamics showcase faster transmission rates than dengue occurring deeper inland. Our results also demonstrate that extreme weather events play a role in the acceleration of dengue transmission along coastlines. Given the similarities between multiple mosquito-borne diseases, it is important to consider this trend for other diseases such as Zika and chikungunya.

Dengue transmission dynamics along coasts are characterized by non-linear dynamics with strong seasonality, multi-annual oscillations, and non-stationary temporal variation (49). Seasonal and muti-annual cycles are fluctuating more unpredictably in the face of climate change, and more irregular intervals of outbreaks are increasingly observed along coastlines (50).

### 4.4 Lag times and dengue transmission dynamics

The time lag or delayed effect of the climatic variables on dengue counts in our model can be reasoned by the factors that indirectly influence dengue counts and transmission. This can happen through effects on vector and virus life-cycle dynamics. The lag is anticipated to be a shorter duration for minimum temperatures that are usually related to the mortality of an adult mosquito, and humidity plays a role in influencing this (29). On the other hand, the lag showed that it would be longer for higher relative water temperatures. Our model shows that the average temperatures, on the other hand, will take longer to influence dengue counts because it is involved in all biological cycles of *Ae. aegypti* (3).

### 4.5 Other factors associated with dengue transmission

There are other factors that must be considered in context of these results, such as socio-economic and demographic factors associated with dengue transmission, as well as the relative contribution of these factors based on scale and geographic topography. These include mosquito management, screen dwellings, the use of insect repellants, protection and prevention programs, bed nets, etc., as well as garden accoutrements, storing water in open containers, etc. These are outside of the scope of our current project, but models should take such data into account.

## Data Availability

The authors will make fully available and without restriction all data underlying our findings. https://github.com/KrtiT/dengue-ms.

## Acknowledgements

We would like to thank the J. William Fulbright Foreign Scholarship Board for funding a year’s worth of unique in-person, on-the-ground field work and data collection that made this entire project possible. We would like to thank both the United States Government and the Government of India for their approval and support for this project. We would also like to give a huge thank you to the Visakha Society for the Protection and Care of Animals (VSPCA), and more specifically, Pradeep Nath, for the home institution that was provided to the lead author while the work was being conducted in India. VSPCA was instrumental in supporting this work and this work would not be possible without them. We would also like to thank the Gandhi Institute of Technology and Management (GITAM) University for the incredible support during this program. The lead author was able to serve as a faculty at GITAM during her time in Andhra Pradesh, and GITAM provided foundational support and guidance for this project. The authors would like to especially thank Dr. Nalini Bikkina, Dr. Divakar Allavarapu, and Dr. Leben Johnson. The authors would further like to take a moment to honor Dr. Leben Johnson, whose life was heartbreakingly taken by COVID-19. We would like to thank the GITAM students whom the lead author taught, for their initial analyses on these data: Amman Naidu, Abhigna Bathina, Balaji Appari, and Kumari Gunuru. The authors would also like to thank the countless institutions that helped them piece this work together: the Andhra Pradesh Forest Department, the Water Resource Board of Andhra Pradesh, the King George Hospital of Visakhapatnam, the Indian Meteorological Department, the UNDP-GoI (USAID) project, and Andhra University, specifically Dr. Professor C. Manjulatha, for her guidance and mentorship. The authors would also like to thank Oxford University, where the lead author sought initial guidance; and Stanford University, where the lead author had a home through which to finalize this research. We would also like to thank the VNU-HCM High School for the Gifted for supporting the second author in his contributions to this work.

## Author Contributions

Conceptualization, K.T.; Data Curation, K.T.; Formal Analysis, M.P.Q. and K.T.; Funding Acquisition, K.T.; Investigation, K.T.; Methodology, K.T. and M.P.Q.; Project Administration, K.T.; Resources, K.T.; Software, K.T. and M.P.Q.; Supervision, K.T.; Validation, M.P.Q. and K.T.; Visualization, M.P.Q. and K.T.; Writing – Original Draft Preparation, K.T.; Writing – Analyses, M.P.Q.; Writing – Review and Editing, K.T; Final Reviews, K.T. and M.P.Q.

All authors have read and agreed to the published version of the manuscript.

## Funding

This research received funding from the Fulbright Program Research Fellowship on behalf of the J. William Fulbright Foreign Scholarship Board.

## Conflicts of Interest

The authors declare no conflict of interest.

## Notes

### Competing Interest Statement

The authors have declared no competing interest.

### Author Declarations

1. https://pcb.ap.gov.in/UI/a_water_quality_monitoring.aspx 2. https://knoema.com/atlas/India/Andhra-Pradesh/topics/Health/Desease-Statistics/Number-of-dengue-cases 3. https://nvbdcp.gov.in/index4.php?lang=1&level=0&linkid=431&lid=3715 4. https://dataful.factly.in/datasets/4125/ 5. https://www.researchgate.net/publication/362720379_Dengue_Fever_Prediction_using_Machine_Learning_Analytics The Dakshin Foundation Ethics Committee waived ethical approval.

